# Early prediction of *Mycobacterium tuberculosis* transmission clusters using supervised learning models

**DOI:** 10.1101/2024.04.16.24305900

**Authors:** Omid Gheysar Gharamaleki, Caroline Colijn, Inna Sekirov, James C Johnston, Benjamin Sobkowiak

## Abstract

Identifying individuals with tuberculosis with a high risk of onward transmission can guide disease prevention and public health strategies. Here, we train classification models to predict the first sampled isolates in *Mycobacterium tuberculosis* transmission clusters from demographic and disease data. We find that supervised learning models, in particular balanced random forests, can be used to develop predictive models that discriminate between individuals with TB that are more likely to form transmission clusters and individuals that are likely not to transmit further, with good model performance and AUCs of ≥ 0.75. We also identified the most important patient and disease characteristics in the best performing classification model, including patient demographics, site of infection, TB lineage, and age at diagnosis. This framework can be used to develop predictive tools for the early assessment of a patient’s transmission risk to prioritise individuals for enhanced follow-up with the aim of reducing further transmission.

## Introduction

Tuberculosis (TB) remains a major global health concern, causing around 1.3 million deaths in 2022 ^1^. The World Health Organization’s “End TB Strategy” sets out aims to eradicate the global TB epidemic by 2035; unfortunately, many countries are currently falling behind targets ^2^. The early diagnosis of people with infectious tuberculosis (TB) disease is key to reducing transmission ^3^. Prioritizing interventions for individuals and groups with a high risk of onward transmission can maximise the impact and cost-effectiveness of public health strategies, such as active case finding and TB preventative therapy, to reduce the spread of disease and prevent outbreaks.

Successful transmission of TB from an infected individual to a contact can be a complex dynamic dependent on the host, pathogen, and environment. Factors associated with the bacteria itself, such as the MTBC lineage, antimicrobial resistance, and the presence of mutations associated with virulence and transmissibility can all contribute to the increased transmission of certain strains ^4–6^. Additionally, environmental and socio-economic conditions can increase the risk of exposure to TB and play a role in transmission ^7^, particularly in low-burden settings where the likelihood of incidental contact with infectious individuals is low.

Identifying the characteristics that are most associated with increased transmission within a population can highlight potential individuals or groups that may transmit onwards and cause outbreaks. Supervised learning can find patterns in complex datasets and build models to accurately predict outcomes in new data. This represents an opportunity to use these approaches in combination with retrospective analysis of past outbreaks to develop predictive models of TB transmission risk from patient and disease characteristics. Developing tools that stratify a patient’s risk of TB transmission using information available to clinicians soon after diagnosis can help to prioritise individuals with an increased risk of transmission for timely interventions and reduce the chance of further infections.

Here, we develop predictive models to discriminate between individuals that with TB that went on to become index cases in transmission clusters and those that did not transmit. We tested multiple supervised learning models using a real-world dataset of *Mycobacterium tuberculosis* (Mtb) whole genome sequence (WGS) data from British Columbia, Canada, a low incidence region of ∼6/100,000 population ^8^. We assess machine learning, deep learning, and logistic regression approaches to classify the first two isolates of transmission clusters using patient demographic and disease characteristics. In addition, we assess the performance of these models when trained to discriminate between the first isolates in larger TB clusters (≥ 3 individuals) and those in non-growing clusters to identify potential infections that contribute most to local transmission and outbreaks.

## Results

### Mycobacterium tuberculosis transmission clusters from British Columbia

A total of 2,575 persons with culture-positive TB were identified in British Columbia, Canada between 2005 and 2015 for who demographic data and a bacterial isolate were available. Mtb whole genome sequence data were obtained for 1,329 isolates collected between 2005 and 2014 that shared a MIRU-VNTR genotyping pattern with at least one other isolate; all isolates with a unique MIRU-VNTR pattern in this period were not sent for further sequencing and were coded as non-clustered in this study. WGS data were available for all 236 TB culture-positive individuals in 2015.

We found that 656/2575 (25.5%) isolates clustered with at least one other isolate using a 12 SNP threshold. This was lower than previous estimates in the province that were calculated using lower resolution genotyping methods (MIRU-VNTR) ^9^. There were 112 transmission clusters overall that ranged in size between 2 and 82 isolates; of these 65/112 (58%) clusters contained two isolates and 17 clusters were of size ≥ 5. Varying the pairwise SNP distance threshold to link isolates did not change the size or composition of clusters significantly. At a 5 SNP threshold, 107 clusters were identified with 598/2575 (23.2%) of isolates in clusters and at a 20 SNP threshold there were 127 clusters with 703/2575 (27.3%) of isolates in clusters; both thresholds identified the same largest cluster comprising the same 82 isolates.

### Classification models

The performance of six supervised learning models (balanced random forest, balanced bagging, balanced logistic regression, LightGBM, TabNet, and a neural network) was evaluated in four classification tasks. The tasks were: **A**) discriminating between the first two isolates by collection date in transmission clusters against non-clustered isolates, **B**) discriminating between the first two isolates by collection date in transmission clusters and all other collected isolates (both non-clustered isolates and clustered isolates that were collected after the first two isolates), **C**) discriminating between the first two isolates by collection date in larger transmission clusters (N ≥ 3) and both non-clustered and clustered isolates in clusters of just two individuals, and **D**) discriminating between the first two isolates by collection date in larger transmission clusters (N ≥ 3) and isolates in clusters of two individuals. Mtb isolates collected between 2005 to 2011 were included as training data and isolates collected in 2012 and 2013 as the test dataset.

**Figure 1** shows ROC curves and AUC values for each tested approach in four classification tasks. We found that most models were able to predict the earliest two isolates in transmission clusters in the test data against non-clustered isolates (task A) with good performance (AUC > 0.7). The best performing models for this task, balanced random forest, LightGBM, and the neural network, had an AUC of 0.81, showing strong discrimination (**Figure 1A**). However, this good model performance may have been driven by the models correctly discriminating simply between hosts belonging to a transmission cluster and those that were non-clustered. Therefore, we next assessed the accuracy in discriminating between the earliest two isolates in transmission clusters in a dataset that included both non-clustered isolates and all other clustered isolates with collection dates later than the first two isolates (task B). Again, the balanced random forest model achieved a good discriminatory power with an AUC of 0.75, followed by the balanced logistic regression approach with an AUC of 0.72 (**Figure 1B**).

**Figure 1.**
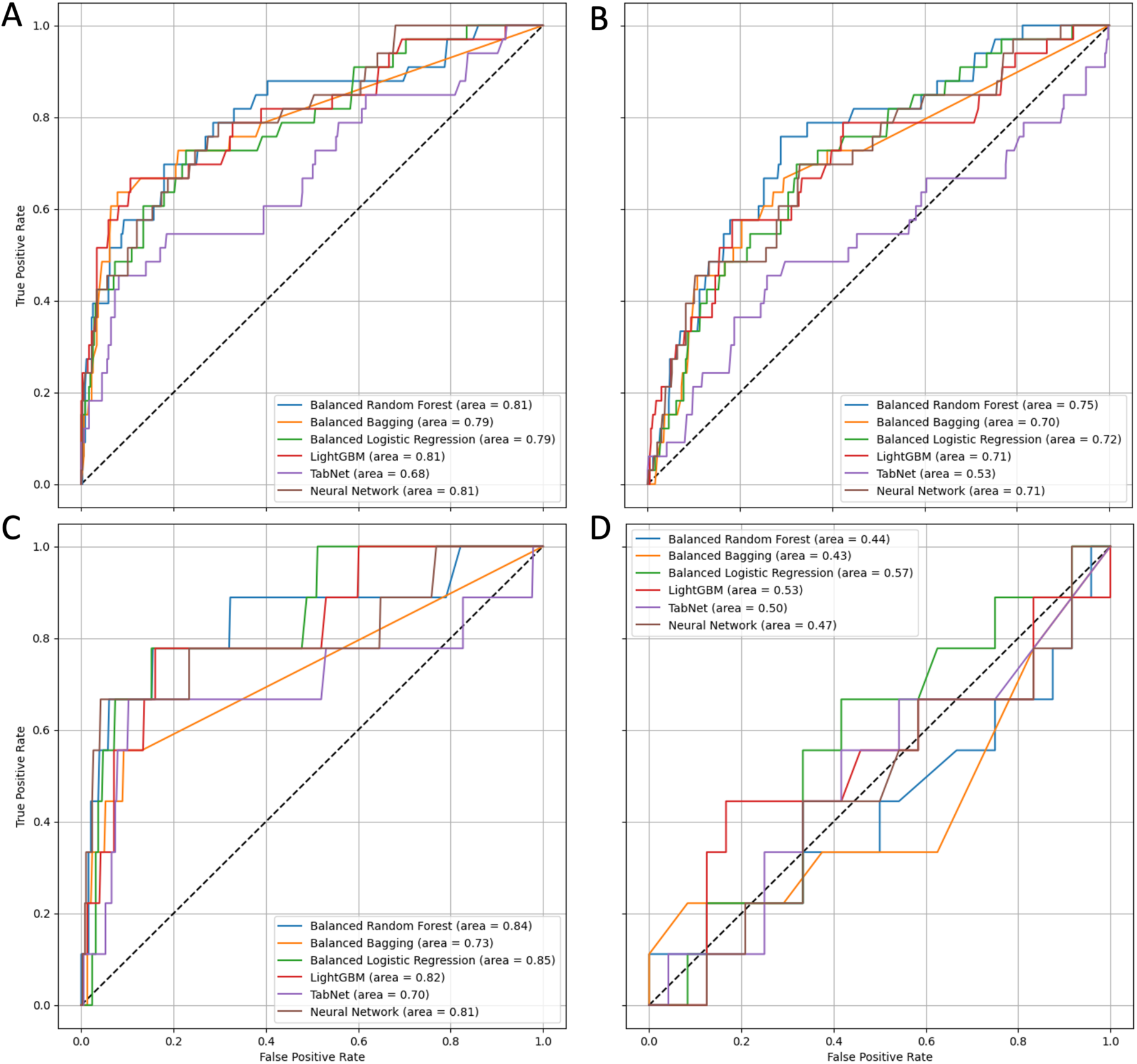
ROC curves and AUC scores for the four classification tasks. A) discriminating between the earliest two clustered isolates and non-clustered isolates, B) discriminating between the earliest two clustered isolates and all other isolates, C) discriminating between the earliest two isolates in clusters of size ≥ 3 and both non-clustered isolates and isolates in clusters of two individuals, and D) discriminating between the earliest two isolates in clusters of size > 2 vs isolates in clusters size = 2.

We next aimed to classify the earliest isolates in larger transmission clusters (N > 2) from a dataset also containing non-clustered isolates and isolates in smaller clusters of two individuals to determine whether there were characteristics specific to the earliest isolates in large clusters (task C). We found that balanced logistic regression achieved the best performance (AUC = 0.85), followed by balance random forest (AUC = 0.84) (**Figure 1C**). Unfortunately, none of the tested models achieved good discrimination when predicting the earliest two isolates in these larger clusters (N > 2) against only isolates from clusters of two individuals (task D) (**Figure 1D**). In this task, the training and test datasets were significantly smaller than tasks A to C, with the test data containing only 34 isolates compared to 391 – 474 in tasks A to C and the training dataset of 164 isolates compared to 1641 – 1341 in tasks A to C.

We identified the patient-level demographic and disease characteristics that contributed most towards the classification in our best performing model, balanced random forest, using feature selection for tasks A to C. We found that patient demographics, site of infection, TB major lineage, and patient age were the features that were the most important to the classification models in predicting the earliest two isolates in transmission clusters for tasks A to C. In addition, the detection method of “symptoms compatible with TB” was of high importance in task B. The full feature rankings for tasks A to C are shown in **Figure S1**.

Next, partial dependence plots were calculated to determine whether specific features had a positive or negative impact on the prediction of the positive response variable (**Figure 2**). We found that Canadian-born individuals were more likely to represent the earliest two isolates in all transmission clusters in tasks A and B, and the earliest two isolates in larger transmission clusters in task C. Individuals with pulmonary TB, symptoms compatible with TB, and carrying a TB strain belonging to the Euro-American lineage were also more likely to be the earliest isolates in clusters in these tasks. Conversely, foreign-born individuals, those aged over 60, and individuals carrying an Indo-American lineage TB strain were negatively associated with a positive response variable and thus were less likely to represent the earliest isolates in clusters in tasks A – C.

**Figure 2.**
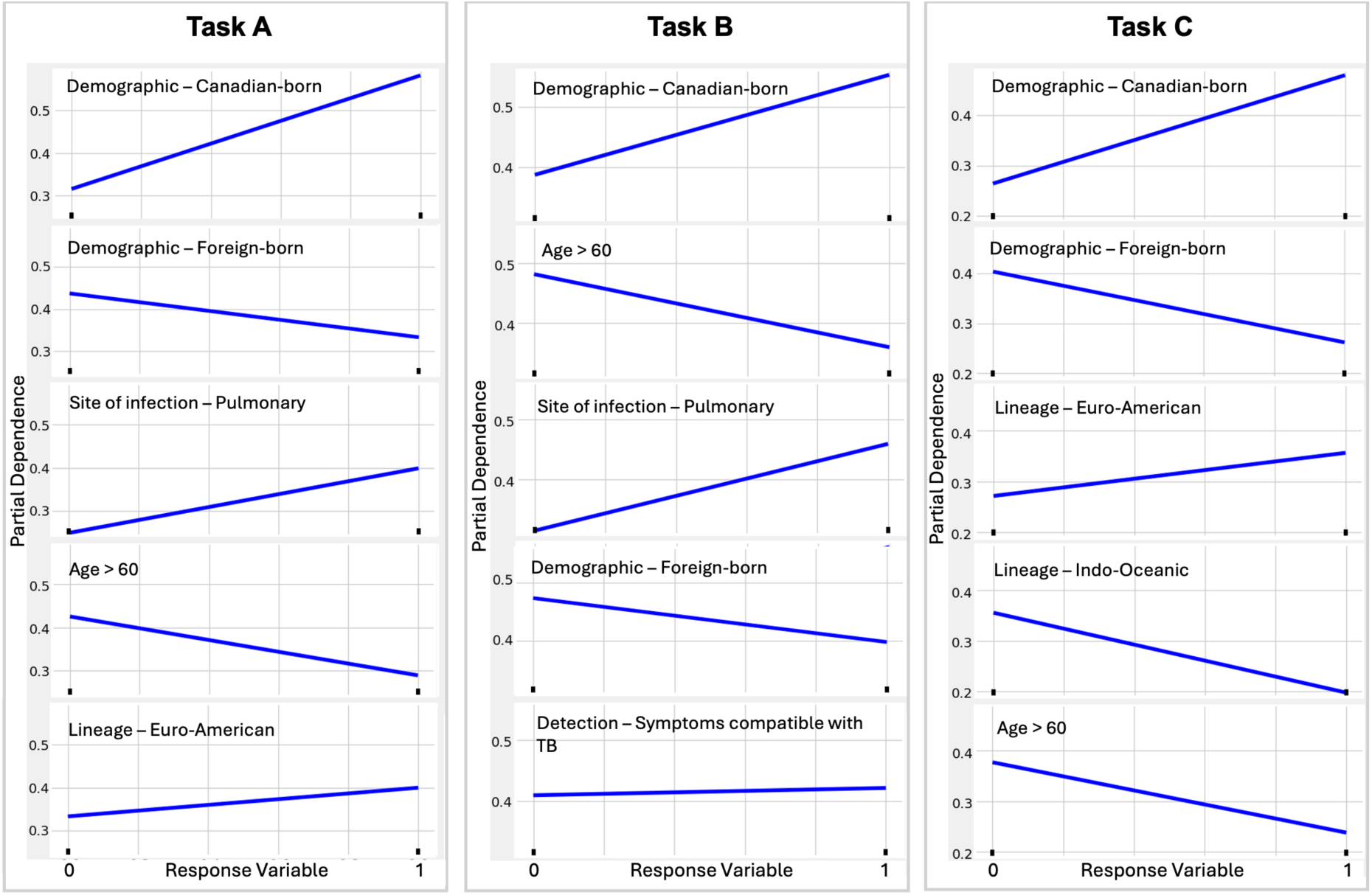
Partial dependence plots of the five patient and disease characteristics with the highest feature importance in the balanced random forest model for tasks A to C. Positive lines represents an increased effect of the named feature on the positive response variable (X axis 0 to 1) for each task.

### Sensitivity analysis

Finally, we found that the good performance of the random forest models in the main analysis was maintained in a sensitivity analysis. **Figure S2** shows the results of the balanced random forest model for classification tasks A, B, and C, with 100 random partitions of the complete 2005 to 2013 dataset using a ratio of 80:20 training to test data. This model performed similarly when trained on these data compared to the original analysis using the 2005 – 2011 training data and 2012 - 2013 test data (AUC 0.75 – 0.81 vs median AUCs 0.73 – 0.90). We also tested the balanced random forest model on tasks A, B, and C when the response variable was randomly assigned (permuted); in this case we would expect models not to perform well as there is no signal to be found. Indeed, the model achieved poor discrimination in all tasks (**Figure S3**), supporting our results that we can predict the earliest isolates in transmission clusters using specific characteristics in these individuals.

## Discussion

Here, we have used patient demographics and disease characteristics to predict which individuals with TB likely go on to form transmission clusters. Using samples collected between 2005 and 2011, we were able to train classification models to predict which patients go on to form transmission clusters in 2012 and 2013 and those that would likely not lead to further transmission events. Recently, machine learning has been used in TB research to improve disease diagnosis, for example the automated detection of lesions on lung x-rays ^10^. Our results suggest that we can use classification models, including machine learning approaches, to classify individuals with TB that represent a higher or lower risk of further transmission through shared demographic and disease features. Furthermore, this work demonstrates the utility of supervised learning models for developing clinically informative tools that can be scaled to incorporate large, complex datasets.

Crucially, we were also able to predict the first two isolates in transmission clusters of all sizes with good discriminatory power using the balanced random forest model. We could also classify isolates that would form larger clusters (three or more isolates) against non-clustered isolates and isolates in clusters of just two individuals. While most clusters beginning in 2012 and 2013 contained just two hosts, there was evidence of further transmission between 2012 and 2015 from 27% of these clusters. The individuals representing the earliest diagnosed infections in the transmission clusters could have been prioritised for investigation using the models presented here to reduce the chance of these clusters growing. The modelling framework presented here can be used to direct epidemiological investigations, which can be costly and labour intensive, and highlight individuals to prioritise for follow-up that may have a higher risk of initiating secondary infections and larger transmission clusters.

There were differences in the model performance of the tested approaches, and we found that the balanced random forest model obtained the best performance overall. The AUC values of the classification models reported here, and the models themselves, are likely to be specific to our TB population, which is in a low-incidence region with high active case finding ^11^. Further limitations include the absence of sputum smear microscopy and drug use disorder data in our analysis, which was not available for a large proportion of individuals, along with limited follow-up time to fully characterize transmission. These factors have previously been reported to increase the risk of TB transmission ^7,9^ and inclusion of these factors would likely impact the performance of the tested classification models. Additionally, the transmission clusters used here were somewhat simple in their construction, with collection dates used to identify individuals that represented the first two hosts in transmission clusters and equal probability of onward transmission from these individuals. The models presented here may be refined by using well-characterized symptom onset times to characterize the isolates that would have become infectious first and reconstructing full transmission networks to infer which individuals likely led to secondary infections later in the transmission cluster.

In task D we were not able to discriminate between the earliest two isolates in larger clusters and those in smaller clusters of only two isolates that did not appear to grow. Both the test and training datasets in this task contained very few individuals and further work would be required to determine whether it is possible to use these models to identify the earliest individuals with TB specifically in growing clusters. This may involve training classification models on larger datasets with more transmission clusters or by including the pairwise covariates between isolates, used previously to predict clustering of un-sequenced individuals with TB using demographic and clinical data ^12^, to increase the information available to train these models.

Nevertheless, key characteristics were identified that were important features used by the classification models to discriminate between the earliest isolates in clusters and both isolates appearing later in clusters and non-clustered isolates. Previous work has identified disease characteristics correlated with Mtb transmission in multiple settings (e.g., ^6,13^), as well as developed complex multilevel models of individual risk prediction for TB ^14^. These studies found evidence of patient and pathogen characteristics that were significantly associated with recent transmission, including TB lineage and age at diagnosis. In our analysis, the most important feature was found to be the patient demographic, with Canadian-born individuals more likely than foreign-born individuals to represent the earliest diagnosed hosts in transmission clusters. Pulmonary TB, previously reported to be associated with an increased TB transmission risk ^15,16^, was also a key feature in tasks A and B of our analysis.

TB lineage was found to be important and though balanced random forest models inherently evaluate the importance of variables independently by averaging across multiple decision trees built from subsets of the features, a much higher proportion of Canadian-born individuals harbored Euro-American lineage TB strains than foreign-born individuals (82% vs 21%) and thus these features were likely to be correlated. Finally, we found patient age to have an impact on classification and patients over the age of 60 had a reduced likelihood of being the earliest isolates in cluster. While the prevalence of TB disease can be high in the elderly through reactivation of latent TB ^17^, the decreased risk of onward transmission shown here in this group may be due to population-specific factors such as social mixing patterns.

In conclusion, we have used classification models to predict the earliest TB isolates in clusters of recent transmission with respect to both host and disease characteristics. This work establishes an easy to implement method to link patient-level correlates of transmission with predictive models to identify persons with TB that may have an increased risk of onward transmission and cluster growth.

## Methods

TB culture-positive individuals diagnosed in BC by the Public Health Laboratory (PHL) of the BC Centre for Disease Control (BCCDC) between 2005 and 2015 were included in this study, for which demographic and clinical data were collected as part of routine clinical investigation. Culture-positive individuals represent approximately 80% of all TB diagnoses in the province ^9^. Sample preparation and DNA extraction was carried out at the British Columbia Public Health Laboratory (BCCDC PHL), as described previously ^9^. WGS data was obtained from individuals sampled between 2005 and 2014 whose TB isolate shared a MIRU-VNTR pattern with at least one other isolate. All isolates collected in 2015 underwent whole genome sequencing. Sequencing and bioinformatic analysis has been described elsewhere ^18^.

Putative clusters representing recent transmission were identified by linking isolates with a maximum pairwise distance of 12 SNPs ^19^. For the classification models, we partitioned the data into a training dataset of isolates collected between 2005 and 2011 and a test dataset of isolates collected between 2012 and 2013. Isolates collected in the last two years of the study were not included in the classification tasks as onward transmission may have occurred after the study period. However, isolates collected between 2005 and 2013 that clustered only with isolates collected in 2014 and 2015 were coded as clustered. Predictor variables were one-hot encoded ^20^ and included demographic data (e.g., sex, age group, and demographics by country of birth) and clinical information (e.g., site of infection and antimicrobial susceptibility) (**Table S1**).

We trained models to carry out four different classification tasks using the 2005 to 2011 training dataset and tested model performance on the 2012 and 2013 test data, with a binary response variable in all instances. In the four classification tasks, the response variable coded as follows: A) a value of 1 for the earliest two isolates in any transmission cluster and 0 for all non-clustered isolates, B) a value of 1 for the earliest two isolates in any transmission cluster and 0 for all other isolates (all non-clustered isolates and all isolates with later collection dates in clusters), C) a value of 1 for the earliest two isolates in transmission clusters of size greater than two and 0 for all non-clustered isolates and isolates in clusters of two, and D) a value of 1 for the earliest two isolates in transmission clusters of size greater than two and 0 for isolates only in clusters of size two.

We tested the performance of multiple classification models, including machine learning (LightGBM, balanced random forest, and balanced bagging), deep learning (Tabnet) models and balanced logistic regression, along with designing a neural network. For the best performing model, the feature importance and effect of the feature on the response variable was calculated to show whether each predictor variable has a positive or negative relationship. All models were run in Python 3.0.

Finally, we carried out two sensitivity analyses using the best performing model overall. Firstly, the training and test data were combined and randomly partitioned into new training and test datasets with an 80:20 ratio to determine whether the classification tasks were influenced by the partitioning of the data by date rather than the predictor variables. Secondly, the response variable in the training data was randomly assigned to isolates, keeping the same proportion of 1s and 0s as in the real data, and all classification tasks re-run on the test dataset. This was to compare our results to the models when trained on training data with a randomly assigned response. All sensitivity analyses were repeated 100 times.

## Data Availability

The whole genome sequence data analyzed in the current study are available from the European Nucleotide Archive (ENA) Project number PRJNA413593.

## Author contributions

B.S., J.C.J., and C.C. conceived and designed the study, J.C.J. and I.S. acquired the data, O.G.G. performed the main analysis under the supervision of B.S., O.G.G., B.S., J.C.J., and C.C. interpreted the results, O.G.G. and B.S. prepared the draft with substantial input from J.C.J. and C.C. All authors revised the manuscript and approved the submitted version.

## Data availability statement

The whole genome sequence data analyzed in the current study are available from the European Nucleotide Archive (ENA) Project number PRJNA413593. The code to run the analysis can be found on GitHub (github.com/bensobkowiak/TBclusterClassification).

## Additional information

### Ethics declarations

Ethics were obtained from the University of British Columbia (certificate H12-00910) and informed consent for participation in the study was not required, as determined by institutional REB review.

---

**Supplementary table S1.**
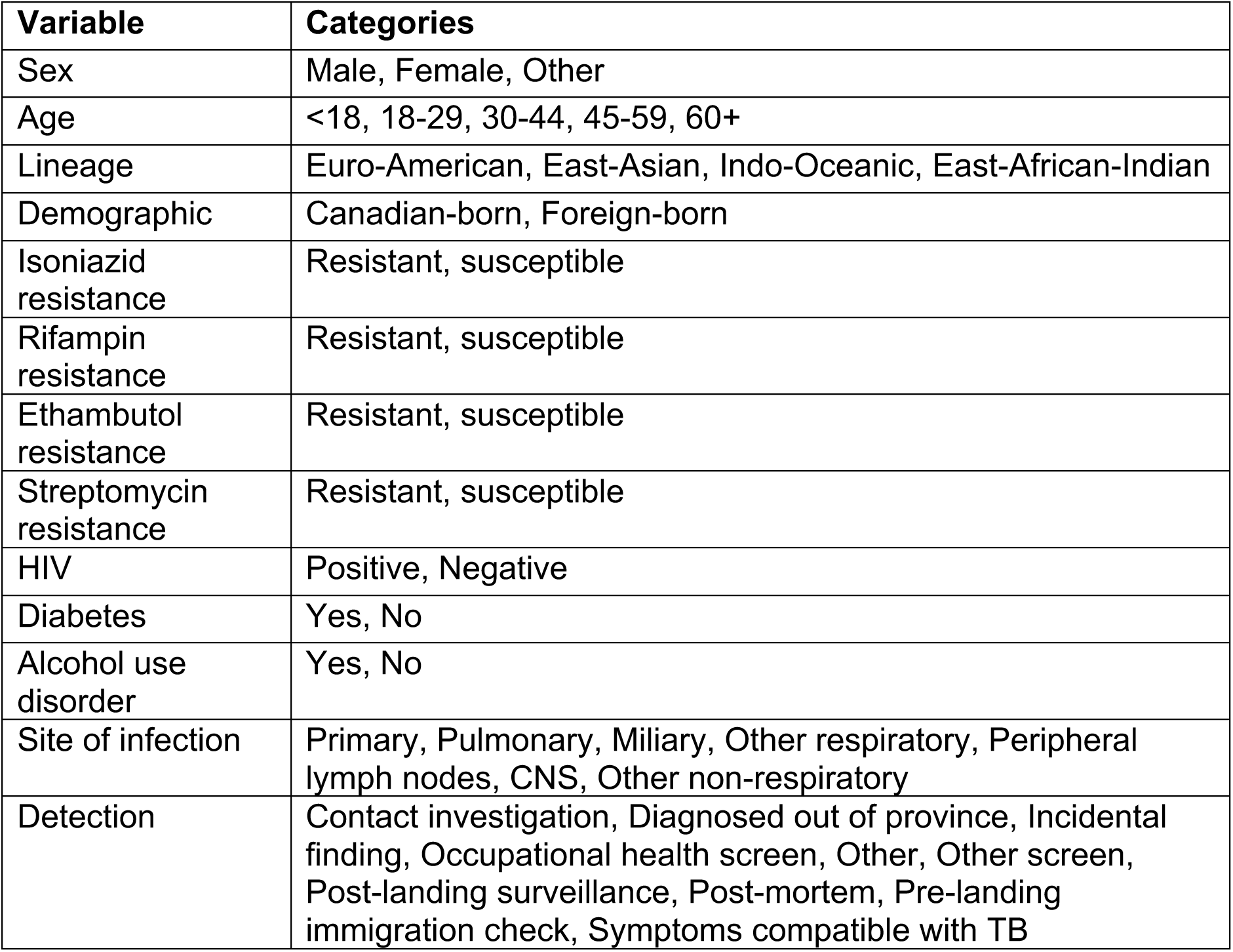
Demographic and disease patient metadata included as independent variables for classification tasks.

**Supplementary figure S1.**
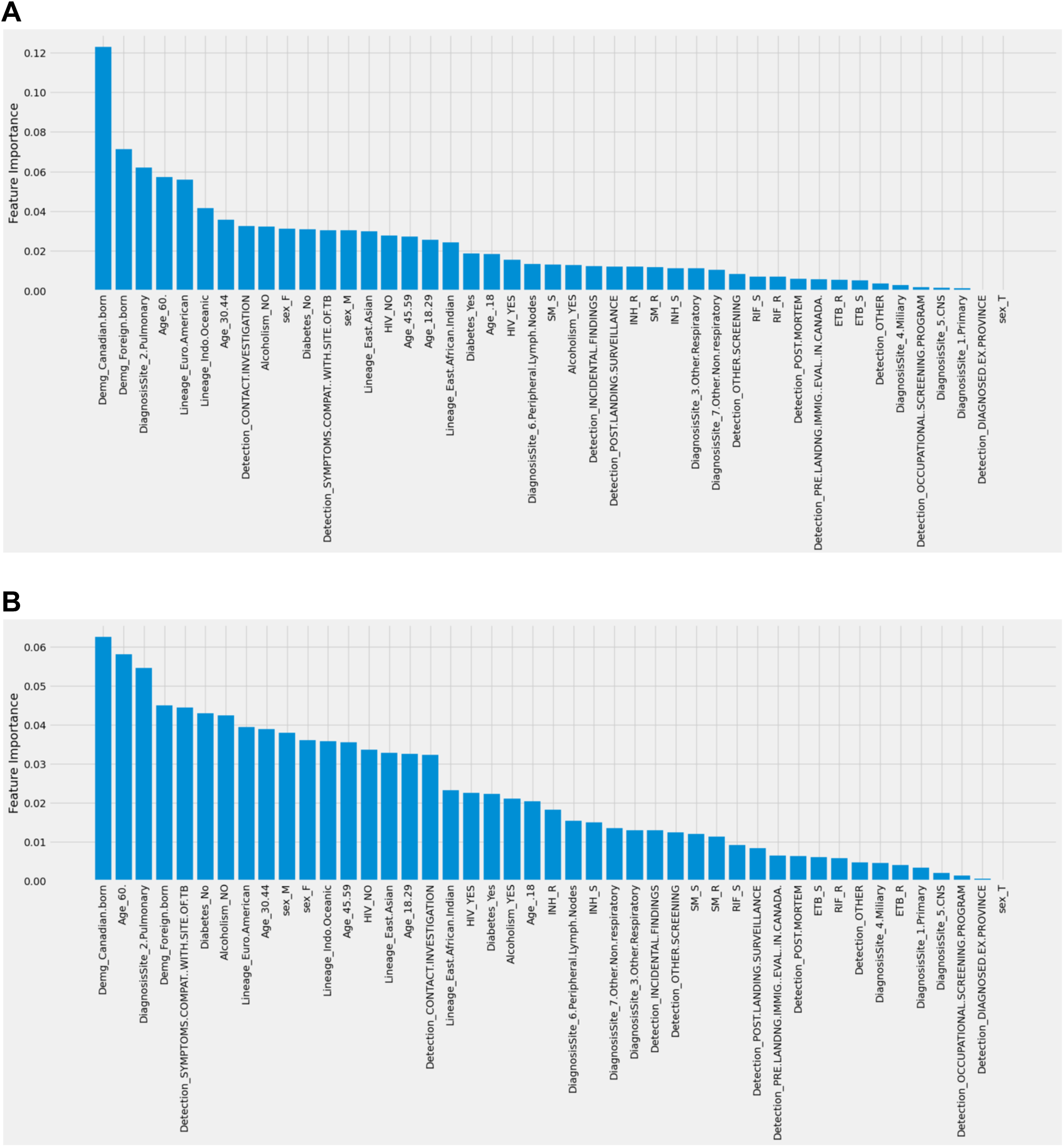
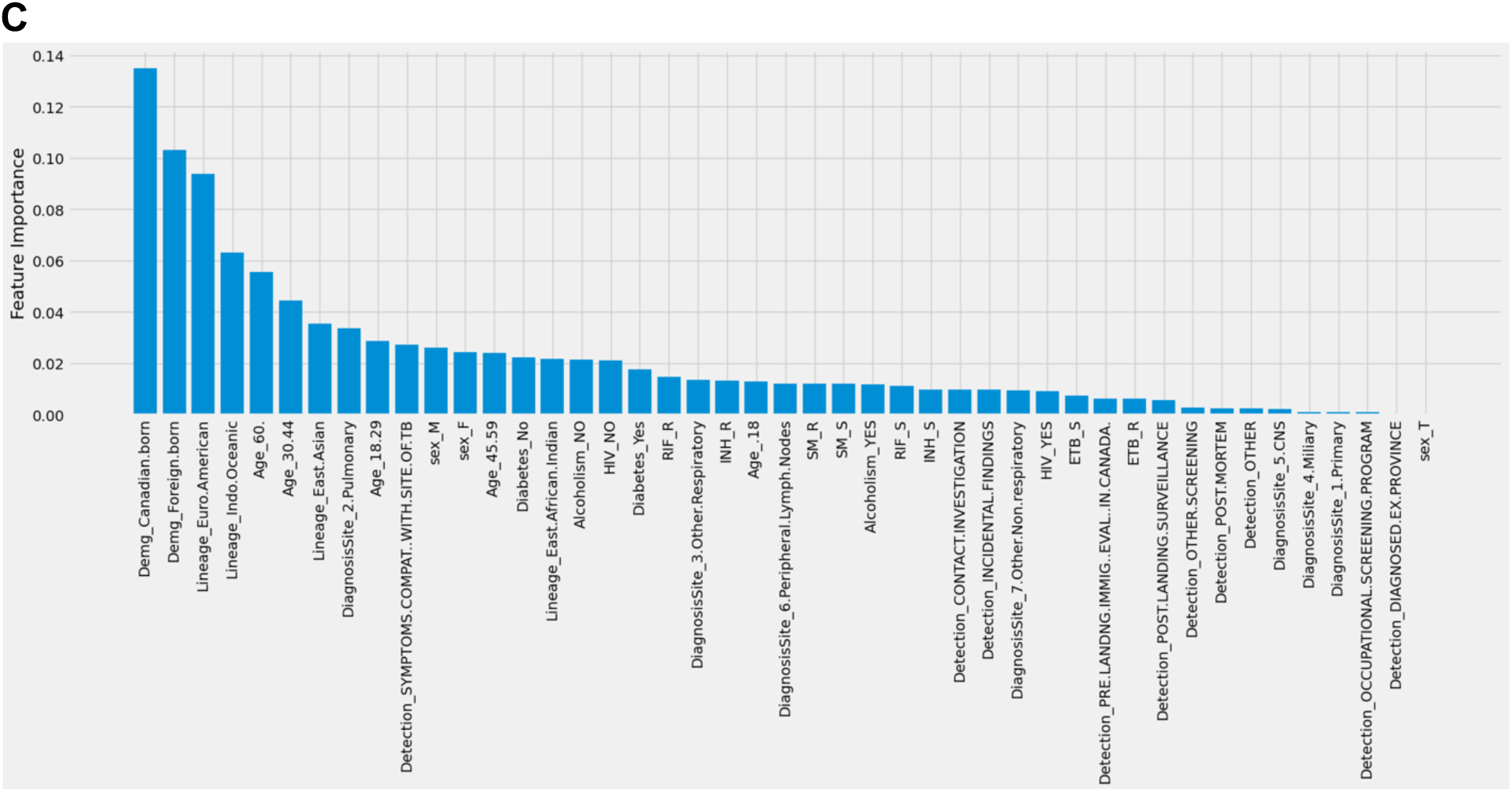
Variable importance for classification using balanced random forest when classifying A) clustered isolates from all data, B) the earliest two isolates in a cluster from non-clustered and earliest clustered isolates, and C) the earliest two isolates in a cluster from all data.

**Supplementary Figure S2.**
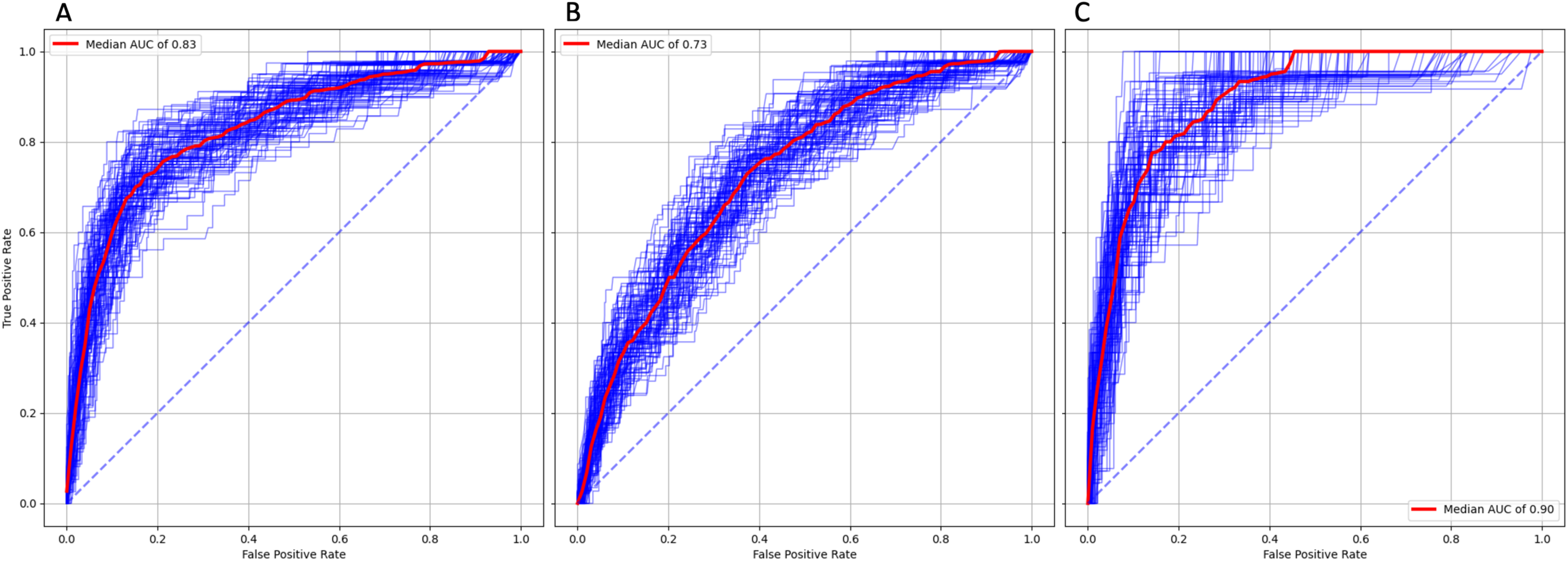
ROC curves and AUC scores for 100 repeats of the balanced random forest model when randomly partitioning the full *Mtb* dataset between 2005 – 2013 into training and test data with an 80:20 ratio. A) Predicting the first two clustered isolates vs non-clustered isolates, B) predicting the first two clustered isolates vs all other isolates, and C) predicting the first two isolates in clusters of size >2 vs non-clustered isolates and isolates in clusters of size = 2.

**Supplementary Figure 3.**
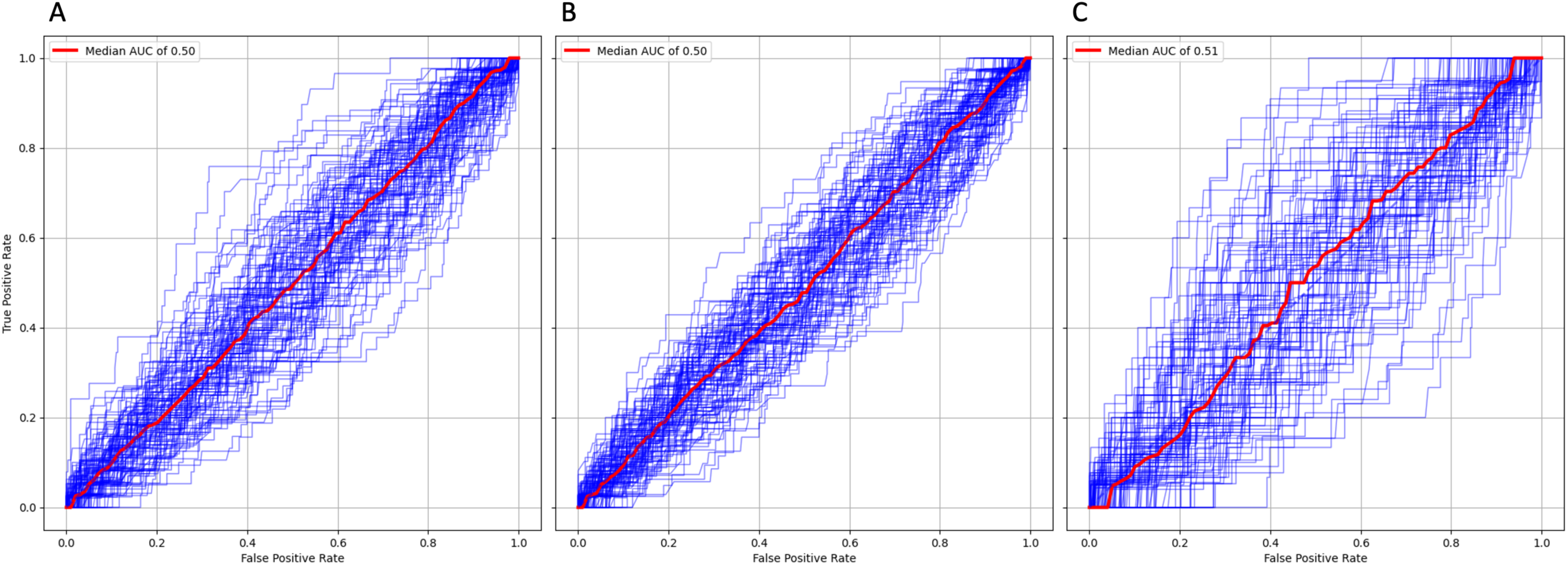
ROC curves and AUC scores for 100 repeats of the balanced random forest model when randomly assigning the response variable in the 2005 – 2011 *Mtb* training dataset. A) Predicting the first two clustered isolates vs non-clustered isolates, B) predicting the first two clustered isolates vs all other isolates, and C) predicting the first two isolates in clusters of size >2 vs non-clustered isolates and isolates in clusters of size = 2.

